# Cue-Induced Craving and Negative Emotion Disrupt Response Inhibition in Methamphetamine Use Disorder: Behavioral and fMRI Results from a Mixed Go/No-Go Task

**DOI:** 10.1101/2021.08.24.21262391

**Authors:** Amirhossein Dakhili, Arshiya Sangchooli, Sara Jafakesh, Mehran Zare Bidoki, Ghazaleh Soleimani, Seyed Amir Hossein Batouli, Kamran Kazemi, Ashkan Faghiri, Mohammad Ali Oghabian, Hamed Ekhtiari

**Affiliations:** Neuroimaging and Analysis Group. (NIAG), Research Center for Molecular and Cellular Imaging, Tehran University of Medical Sciences, Tehran Iran; Medical Physics Department, Iran University of Medical Sciences, Tehran, Iran; Iranian National Center for Addiction Studies (INCAS), Tehran University of Medical Science, Tehran, Iran; Department of Electrical and Electronics Engineering, Shiraz University of Technology, Shiraz, Iran; School of Medicine, Shahid-Sadoughi University of Medical Sciences, Yazd, Iran; Department of Biomedical Engineering, Amirkabir University of Technology (Tehran Polytechnic), Tehran, Iran; Medical Physics and Biomedical Engineering Department, Tehran University of Medical Sciences, Tehran Iran; Tri-Institutional Center for Translational Research in Neuroimaging and Data Science (TReNDS), Georgia State University, Georgia Institute of Technology, and Emory University, Atlanta, GA 30303, USA; Laureate Institute for Brain Research (LIBR), Tulsa, OK, USA

**Author notes:** **Corresponding Author:** Hamed Ekhtiari, MD, PhD, Laureate Institute for Brain Research, 6655 South Yale Ave. Tulsa, OK 74136.

**Keywords:** Methamphetamine, fMRI, Cue reactivity, Craving, Response inhibition, Go/No-Go

## Abstract

**Background:** Drug-related cue-reactivity, dysfunctional negative emotion processing, and response-disinhibition constitute three core aspects of methamphetamine use disorder (MUD). These phenomena have been studied independently, but the neuroscientific literature on their interaction in addictive disorders remains scant.

**Methods:** fMRI data were collected from 62 individuals with MUD when responding to the geometric Go or No-Go cues superimposed over blank, neutral, negative-emotional and drug-related background images. Neural correlates of drug and negative-emotional cue-reactivity, response-inhibition, and response-inhibition during drug and negative-emotional blocks were estimated, and methamphetamine cue-reactivity was compared between MUDs and 23 healthy controls (HCs). Relationships between clinical and behavioral characteristics and observed activations were subsequently investigated.

**Results:** MUDs had longer reaction times and more errors in drug and negative-emotional blocks compared to neutral and blank ones. MUDs showed higher drug cue-reactivity than HCs across prefrontal regions, fusiform gyrus, and visual cortices (Z>3.1, p-corrected<0.05). Response-inhibition was associated with activations in the precuneus, inferior parietal lobule, and anterior cingulate, temporal and inferior frontal gyri (Z>3.1, p-corrected<0.05). Response-inhibition in drug cue blocks coincided with higher activations in the visual cortex and lower activations in the paracentral lobule and superior and inferior frontal gyri, while inhibition during negative-emotional blocks led to higher superior parietal, fusiform, and lateral occipital activations (Z>3.1, p-corrected<0.05).

**Conclusion:** Higher visual cortical activations and lower parietal and prefrontal activations during drug-related response-inhibition suggest the down-regulation of inhibitory regions and up-regulation of bottom-up drug cue-reactivity. Our results suggest that drug and negative-emotional cue-reactivity influence response-inhibition, and the study of these interactions may aid mechanistic understandings of addiction and biomarker discovery.

## 1. Introduction

Methamphetamine is the most used amphetamine-type stimulant, with increasing use around the globe (United Nations Office on Drugs and Crime, 2018). The prevalence of methamphetamine use disorder (MUD) in the US has increased by 195% between 2010 to 2018, with a 200% increase in deaths due to overdose from 2011 to 2016, leading some to suggest that it might be responsible for the next substance use crisis in the US (Hedegaard et al., 2018). Considering the current need for more effective interventions, it has been argued that a better understanding of the neurobiology of MUD is necessary and could lead to the discovery of novel treatment targets (Paulus and Stewart, 2020).

There is mounting evidence that people with MUD have a variety of cognitive impairments, such as inhibitory control over substance use (Monterosso et al., 2005; Nestor et al., 2011) a heightened subjective and neurophysiological response to methamphetamine-related cues (Alam Mehrjerdi et al., 2011; Ekhtiari et al., 2020), and dysfunctional responses to negative emotional cues (henceforth referred to as “negative cues”) (May et al., 2020). Recent evidence suggests that these three domains are affected across substance use disorders (SUDs). There is a consensus that the cognitive control, positive valence and negative valence research domain criteria (RDoC) are uniquely crucial in SUDs (Yücel et al., 2019). The impaired response-inhibition and salience attribution (IRISA) model of addiction also proposes that people with SUDs have problems in both cognitive control over substance use and attributing high saliency to substance-related rewards (Goldstein and Volkow, 2002, 2011), and the Addictions Neuroclinical Assessment (ANA) model is based around the three domains of incentive salience, negative emotionality, and executive function (Kwako et al., 2017).

A growing literature has painted a complex pattern of neural activity underlying the dysfunctions in these domains. Impairments of inhibitory control, assessed using paradigms such as the Go/No-GO task (Luijten et al., 2014), are associated with different activation patterns in SUDs compared to healthy controls (HCs) in the supplementary motor area, insula, precuneus, and many temporal and frontal regions (Kelly et al., 2004; Mostofsky et al., 2003; Nestor et al., 2011; Paulus et al., 2005; Simmonds et al., 2008). Drug cue-reactivity, the host of behavioral, physiological and neural responses to drug cues which are associated with a cue-induced craving for addictive substances and subsequent drug use behavior (Ekhtiari et al., 2021), is associated with activations across the frontal, insular, striatal and limbic regions, overlapping those involved in dysfunctional response inhibition (Dean et al., 2019; Grodin et al., 2019; Guterstam et al., 2018; Yin et al., 2012; Zilverstand et al., 2018). Importantly, there is evidence that response-inhibition and drug cue-reactivity can interact (Ames et al., 2014; Stein et al., 2021; van Holst et al., 2012), and some fMRI correlates of drug cue-reactivity, and response-inhibition can predict successfully management of impulses in daily life (Lopez et al., 2014).

Response-inhibition can also interact with reactivity to negative cues and the processing of negative affect (Curci et al., 2013; Cyders and Coskunpinar, 2011; Cyders and Smith, 2008). Impulsive responses when experiencing negative emotion can be referred to as negative urgency, which is correlated with dysfunctional activations in brain regions undergirding the interaction of emotion regulation and cognitive control such as the anterior insula and the orbitofrontal cortex (Johnson et al., 2020), potentially implicating the failure of top-down inhibitory control when processing emotionally salient stimuli (Heatherton and Wagner, 2011). It is well known that negative urgency and its underlying neural dysfunctions are implicated in addictive disorders (Kaiser et al., 2012; Um et al., 2019).

Despite suggestions that drug cue-reactivity and negative emotion processing influence response-inhibition in SUDs, most task-based fMRI studies of SUDs have examined these phenomena separately. Using a novel fMRI task that combines a Go/No-Go task with drug cue-reactivity and negative emotional cue-reactivity paradigms using validated cue databases, this study aims to investigate the interactions of response-inhibition, drug cue-reactivity and negative emotional cue-reactivity and their neural substrates. We hypothesize that individuals with MUD will perform more poorly in the Go/No-Go task when exposed to methamphetamine-related and negative cues than neutral cues. Exposure to these cues will modulate the brain activity associated with response-inhibition.

## 2. Materials and Methods

### 2.1 Participants

Male participants with MUD, aged 20-40 years, were recruited from multiple addiction treatment centers in Tehran, Iran. All participants met the following inclusion criteria: (1) Diagnosis of methamphetamine dependence (for at least six months) according to the Diagnostic Statistical Manual of Mental Disorders, Fourth Edition, Text Revision *(American Psychiatric Association, 2013)*, (2) abstinence from any substance except nicotine for at least a week, confirmed by negative urine drug screening and self-report, (3) right-handedness, as determined using the Edinburgh Handedness Inventory (Oldfield, 1971). Exclusion Criteria were: (1) comorbid axis-I disorders, other than drug dependence (According to DSM-IV-TR), (2) ineligibility for MRI scanning (e.g., metal implants, claustrophobia), (3) head trauma with neurologic sequelae, 4) neurologic disorder which interferes with the research process. Written informed consent was obtained from all participants. A total of 75 MUDs were screened in the laboratory. Thirteen did not meet inclusion criteria, leaving 62 participants who enrolled and completed the entire protocol. Nine MUDs were excluded from data analysis due to high head motion during scanning, leaving a total of 53 MUDs for fMRI analyses.

To validate the drug cue-reactivity aspect of the task, 23 healthy individuals were chosen as HCs. The HCs had no past or current diagnosis of substance use disorder or history of methamphetamine use and otherwise had the same inclusion and exclusion criteria as MUDs.

The study was conducted in accordance with the Declaration of Helsinki. Individuals with MUD were told that the fMRI task could induce methamphetamine craving and they would be asked to remain in the scanning center for an hour to recover. Participants then provided written, informed consent prior to further screening for enrollment. All collected data were anonymized by the data analyst before further analysis. The study protocol was approved by the ethical review board of the Tehran University of Medical Sciences with the approval code 93-02-98-23869.

### 2.2 Measures and Questionnaires

Potential participants were assessed by two clinical psychologists. Several questions and measures were administered before scanning, including demographic information, substance use profile, treatment history, risky behaviors profile (Injection History, Sexual Intercourse, Imprisonment History, Drug Selling History and Fight History), the Barratt Impulsiveness Scale-11 (BIS-11) (Barratt, 1994) and the Depression Anxiety and Stress Scale-21 (DASS-21) (Osman et al., 2012). Before and after scanning, MUDs also rated their drug craving on a 0-100 Visual Analog Scale (VAS) and rated a positive and negative scale (PANAS) (Crawford and Henry, 2004).

### 2.3 Mixed Go/No-Go task

All MUDs and HCs were scanned during four consecutive runs of the mixed Go/No-Go task, separated by resting blocks with a fixation point. Each run consisted of four 36-second blocks, each containing 24 pictures depicting geometric Go/No-Go stimuli superimposed on the background cues. Background images were either blank, neutral cues, negative cues, and drug cues. The order of the blocks was pseudo-randomized in each run. The 24 drug-related cues have been evaluated in previous studies (Ekhtiari et al., 2010) and the neutral and negative cues were selected from the International Affective Picture System database and matched for visual complexity (Lang et al., 1997). Participants viewed each cue only once. Go stimuli were triangles, squares, or diamonds and No-Go stimuli were circles. Of the 24 trials in each block, 6 were No-Go and 18 were Go trials. Each stimulus lasted for 1 second and was followed by a jittered interstimulus interval (generated using a gamma function, mean=.5). Blocks were separated by 18-second fixation periods in which a white cross was shown on a black background. Participants were asked to respond as fast as possible when the Go stimuli were presented and withhold their response when seeing No-Go stimuli. Before the scanning session, participants underwent a training test outside the scanner and were informed that both speed and accuracy are important (Figure 1).

**Figure 1:**
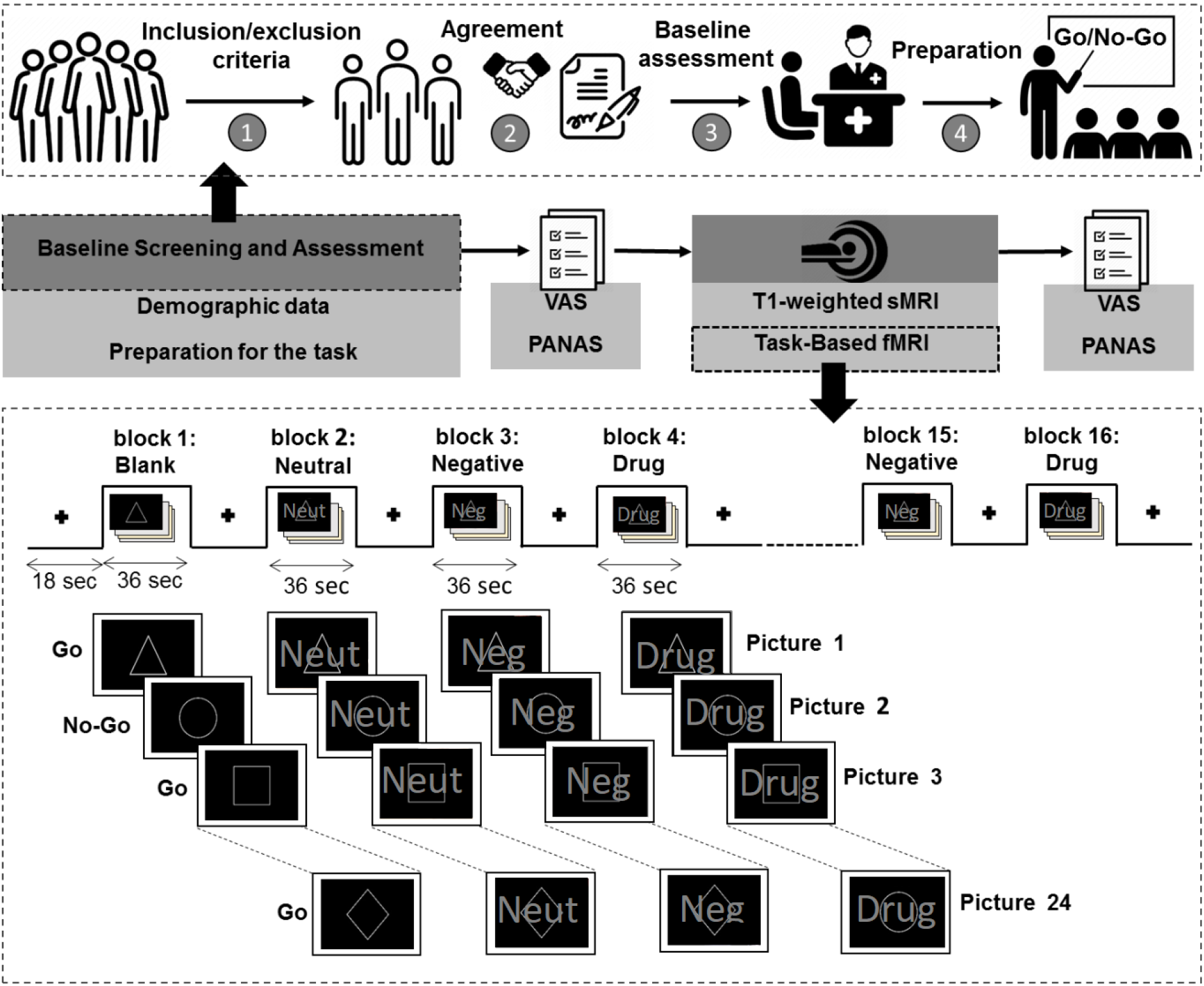
Summary of the study protocol and the mixed Go/No-Go cue-reactivity task. At the baseline screening and assessment, (1) if subjects passed our inclusion/exclusion criteria, they were invited to the center for further assessment. (2) Subjects were asked to sign the agreement. (3) Subjects were interviewed by a clinical psychologist to collect baseline data. (4) Subjects were trained for the Go/NoGo task and prepared for the scanning session. Immediately before and after scanning PANAS and VAS measures were administered to each subject. During the scanning session, high-resolution T1-weighted and task-based fMRI were collected. The bottom part of the figure shows the Go/NoGo fMRI task design. Each of the four runs of the task included four blocks, adding up to 16 blocks overall. In each block, geometric Go and No-Go stimuli were superimposed on either blank, neutral, emotionally negative, or methamphetamine-related cues. The ‘Go’ stimuli were polygonal and the ‘NoGo’ stimulus was a circle. Reaction times were collected during the task for each individual.

### 2.4 Scanning Parameters

Scanning was conducted in a 3.0 Tesla (Siemens, MAGNETOM Trio; Germany) MR-system (Neuroimaging and Analysis Group (NIAG); Imam Khomeini Hospital, Tehran, Iran). Structural T1-weighted images were acquired in a sagittal orientation employing a magnetization-prepared rapid gradient-echo (MP-RAGE) sequence with the following parameters: Repetition Time =1800ms, Echo Time = 3.44ms, field of view (FOV) = 256 cm × 256 cm, flip angle (FA) = 7°, 1mm3 Voxels. Functional MRI data were obtained using a gradient-echo echo-planar imaging (GRE-EPI) sequence with the following parameters: FOV = 192 × 192, FA = 90°, in-plane voxel size 3 m. Each run lasted 13 minutes and 26 seconds.

### 2.5 Pre-processing

fMRI analysis was performed using the fMRI Expert Analysis Tool (FEAT), part of FMRIB’s Software Library version 6.0.3. The preprocessing procedure consisted of: 1) Skull-stripping to remove non-brain tissue from the structural T1-weighted images, using the Brain Extraction Tool (BET) with default values, 2) Removal of first five time points, 3) Motion correction with 6 degrees of freedom (DOF), 4) Interleaved slice-timing correction 5) Spatial smoothing using a Gaussian kernel of FWHM= 5.0 mm, 6) Melodic ICA data exploration to identify remaining data artifacts and to help exploring activation in the data, 7) Multiplicative mean intensity normalization of the volume at each time point, 8) High-pass temporal filtering (Gaussian weighted least-squares straight-line fitting, with Inverse of= 120.0s), 9) Co-registration of the functional images to the self-high resolution using FMRIB’s Linear Image Registration (FLIRT) and Boundary-Based Registration (BBR) cost function, 10) Nonlinear registration of the structural T1 images to the MNI space with 12 degrees of freedom, and 11) Despiking to regress-out the remaining motion effects on fMRI time-series, identified with the DVARS metric given by the FSLMotionOutliers tool. Subjects with a DVARS-threshold>75 on more than 10 time points in a single block were excluded from the analysis.

### 2.6 Statistical Analysis

Anatomic labeling, and locating the activations were performed using the Brainnetome atlas (BNA)(Fan et al., 2016), with a subsequent visual inspection of different activation clusters, overlaid on the T1-weighted image of the MNI152 atlas.

The first-level General Linear Model (GLM) statistical analysis was performed with a Z-threshold of 3.1 and the Cluster Defining Threshold (CDT) method with a corrected p-value<0.05. In order to calculate the average brain activation for each contrast, higher-level analyses were performed using FMRIB’s Local Analysis of Mixed Effects (FLAME) tool (Z-threshold=>3.1, alpha corrected p-value<0.05).

Event-types were specified at the time of indicator onset and the canonical hemodynamic response was used to model the regressors for the conditions of interest. The event types included Blank Successful No-Go (BSNG), Blank Successful Go (BSG), Neutral Successful No-Go (NSNG), Neutral Successful Go (NSG), Negative Successful No-Go (NENG), Negative Successful Go (NESG), Drug Successful No-Go (DSNG), and Drug Successful Go (DSG). Each event type was included as a single regressor. Unsuccessful trials were modeled by a single nuisance regressor in the GLM. Cue-reactivity contrasts were examined as *(Drug>Neutral)* for drug cue-reactivity and *(Negative>Neutral)* for negative emotional cue-reactivity for MUDs and HCs. The response-inhibition contrast was defined as [*(BSNG+NSNG+NESNG+DSNG> BSG+NSG+NESG+DSG)]*. The interaction between cue-reactivity and response-inhibition was modeled with the *[(DSNG>DSG)>(NSNG>NSG)]* contrast for drug and *[(NESNG>NESG)>(NSNG>NSG)]* contrast for negative cues. Six realignment and high-motion parameters based on DVARS metrics were included as nuisance regressors to correct head movement. Finally, a two-sample t-test had been done to compare MUDs and HCs, was calculated

All behavioral and clinical data calculations were performed using R software. Masks and first-level parameter estimates for the activation clusters in each contrast were extracted (R Core Team, 2013), and the correlations of subject parameter estimates and the relevant behavioral and clinical variables, including meth use duration, pre-and post-scanning VAS, pre-and post-scanning negative PANAS, BIS (sum and motor subscale scores), risky behavior history, and commission error rates on the Go/No-Go task. We also compared MUDs with and without drug use in the month before scanning.

## 3. Results

### 3.1 Participant characteristics

Demographic and clinical profiles are summarised in Table 1. Significant differences were found between HCs and MUDs in terms of BIS and DASS scores.

**Table 1.** Characteristics of participants (n=76).

| Variables | MUDs (n=53) | Healthy Controls (n=23) |
| --- | --- | --- |
| **Demographic Variables** |  |  |
| Age (years) | 32.12(5.89) | 31.17(5.69) |
| Education (years) | 10.08(3.07) | 11.91(2.96) |
| **Substance use Patterns** |  |  |
| Total abstinence duration (Day) | 81.6(121.5) | - |
| Total drug abuse duration (months) | 146.3(77.3) | - |
| **Hallucinogens** |  |  |
| Number of users | 4(7%) | - |
| Use frequency | 2.83(7.5) | - |
| **Opioids** |  |  |
| Number of users | 47(88%) | - |
| Use frequency | 22.96(12.6) | - |
| **Cannabis** |  |  |
| Number of users | 34(64%) | - |
| Use frequency | 16.5(14.3) | - |
| **Sedatives** |  |  |
| Number of users | 12(22%) | - |
| Use frequency | 5.6(10.9) | - |
| **Cocaine** |  |  |
| Number of users | 4(7%) | - |
| Use frequency | 2.6(8.1) | - |
| **Alcohol** |  |  |
| Number of users | 22(41%) | - |
| Use frequency | 12(13.8) | - |
| **Treatment History** |  |  |
| Treatment in last month | 1.98(0.12) | - |
| Times of treatment in last year | 2.67(3.60) | - |
| NA participation history | 1.77(0.42) | - |
| NA Duration (Months) | 15.46(25.42) | - |
| **Clinical Scales** |  |  |
| BIS Scores | 75.08(15.11) | 57.4(14.87)* |
| DASS Scores | 27.53(13.60) | 24.4(11.52)* |
| **Risky Behaviors** |  |  |
| Number of risky behaviors | 2.26(1.11) | - |
| Injection History | 10(18%) | - |
| Sexual Intercourse | 52(98%) | - |
| Imprisonment History | 24(45%) | - |
| Drug Selling History | 22(41%) | - |
| Fight History | 14(26%) | - |

**Note:** Values are denoted either as mean (SD) or number(percentage). For each substance category, users are defined as individuals who have ever used a substance belonging to that category for at least one month. "Use frequency" for each substance category is defined as the number of days any substance belonging to that category had been used, during the last month in which substances of that category were used. For individuals who used more than a single substance from one category, the most frequently used substance is chosen as a stand-in for the frequency of using substances from that category. "Risky Behaviors" is a score ranging from one to five, indicating the number of risky behavior domains the participant has engaged in. These include "having ever had illicit sexual intercourse", "having ever been imprisoned", "having ever injected drugs", "having ever sold drugs", and "having gotten into any fights in the last month". Abbreviations: BIS, Barratt Impulsiveness Scale; DASS, Depression Anxiety Stress Scale; VAS, Visual Analog Scale. "Risky Behaviors" is a score ranging from one to five, indicating the number of risky behavior domains the participant has engaged in. These include "having ever had illicit sexual intercourse", "having ever been imprisoned", "having ever injected drugs", "having ever sold drugs", and "having gotten into any fights in the last month".
\*: Data for these cells exist only for 5 Healthy controls.

### 3.2 Behavioural performance on the Go/No-Go task

The percentage of commission errors in neutral (Mean=11.32, SD=10.50), negative (Mean=12.50, SD=11.29) and drug (Mean=13.28, SD=13.89) blocks were significantly higher than in blank blocks (Mean=6.52, SD=7.98). The percentage of omission errors in neutral (Mean=9.93, SD=10.59), negative (Mean=12.42, SD=10.25) and drug (Mean=12.91, SD=11.09) cue blocks were also significantly higher than blank blocks (Mean=3.09, SD=3.79). The percentages of commission and omission errors in drug and negative blocks were also significantly higher than neutral cue blocks. Furthermore, there was no significant difference between the reaction times of MUDs in neutral, negative and drug cue blocks, although reaction time for blank cue blocks was much lower (Mean=674.27, SD=69.61) than neutral (Mean=761.37, SD=60.17), negative (Mean=768.40, SD=54.31) and drug (Mean=757.27, SD=59.46) blocks (Figure 2.a and Table S1). More behavioral outcomes based on the changes in the PANAS and VAS scores can be found in Figure S1.

**Figure 2:**
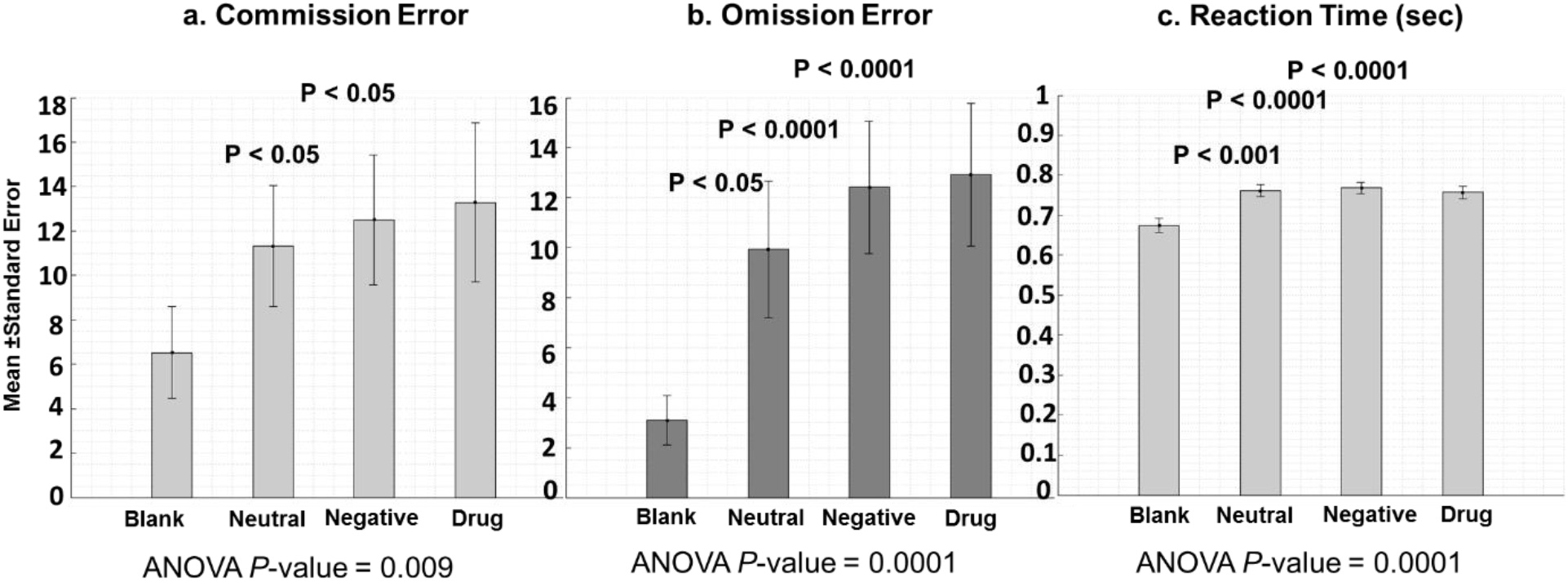
Behavioral performance. Behavioral performance of participants with MUD on the Go/No-Go task across the four different block types. ANOVA *P-*values show significant differences in the three group comparisons. P-values of post-hoc tests (above bar charts) show significant differences in reaction times/ commission error/ omission error rates between different block types. Bars show mean values and error bars represent the standard error of the mean. Omission error is defined as failing to respond to Go trials, and commission error is the failure to inhibit pre-potent responses on No-Go trials.

### 3.3 Drug and negative cue-reactivity

In MUDs, drug cue exposure was associated with higher activity compared to neutral cue exposure in the left lateral superior frontal gyrus (SFG) (4263 voxels, z-max=6.7), left rostroventral inferior temporal gyrus (ITG) (1179 voxels, z-max=6.1), left dorsal (955 voxels, z-max=7.14), caudodorsal (297 voxels, z-max=5.16) and pregenual cingulate gyrus (CG) (221 voxels, z-max=4.86), left ventral caudate basal ganglia (BG) (758 voxels, z-max=5.14), left caudal inferior parietal lobule (IPL) (409 voxels, z-max=5.34) and rostroventral IPL (357 voxels, z-max=4.7), left inferior frontal sulcus (IFS) (406 voxels, z-max=6.14), right lateral occipital cortex (LOcC) (375 voxels, z-max=5.47), left orbital gyrus (ORG) (312 voxels, z-max=5.06), caudal ventrolateral precentral gyrus (PrG) (177 voxels, z-max=4.8), and lower activity compared to neutral cue exposure in the right caudal cuneus (12600 voxels, z-max=7.76), left superior temporal gyri (STG) (planum STG (500 voxels, z-max=5.04), anterior STG (137 voxels, z-max=4.08), and paracentral lobule (PCL) (110 voxels, z-max=4.91) corrected p-value<0.05 (Figure 3.a and Table S2).

**Figure 3:**
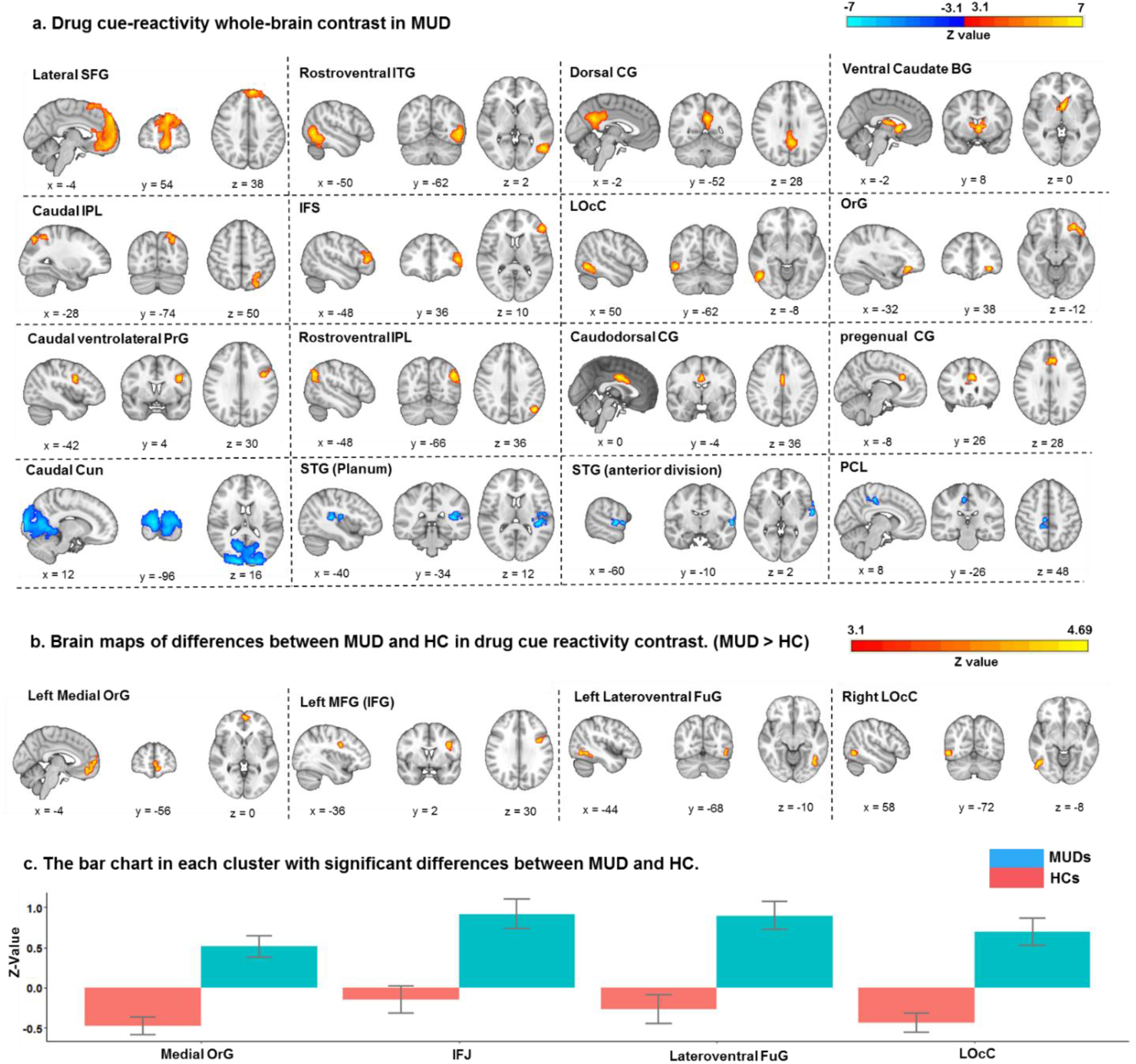
Drug cue reactivity. **a.** Significant clusters in the drug cue-reactivity whole-brain contrast in MUDs surviving the FDR-corrected p<0.05 threshold. Positive z-values (in warm colors) indicate higher voxel activations associated with methamphetamine compared to neutral cues, while negative z-values (in cold colors), indicate the reverse. MNI coordinate of the voxel with the maximal z-value in each cluster is reported bellow sagittal, coronal, and axial views. 16 significant clusters are visualized based on the cluster-defining threshold method and reported based on the Brainnetome atlas parcellation: left lateral superior frontal gyrus (Lateral SFG, 4263 voxels), left rostroventral inferior temporal gyrus (Rostroventral ITG, 1179 voxels), left dorsal cingulate gyrus (Dorsal CG, 995 voxels), left ventral caudate basal ganglia (Ventral Caudate BG, 758 voxels), left caudal inferior parietal lobule (Caudal IPL, 409 voxels), left rostroventral inferior parietal lobule (Rostroventral IPL, 357 voxels), left inferior frontal sulcus (IFS, 406 voxels), right lateral occipital cortex (LOoC, 375 voxels), left orbital gyrus (OrG, 312 voxels), left caudodorsal cingulate gyrus (Caudodorsal CG, 27 voxels), left pregenual cingulate gyrus (Pregenual CG, 221 voxels), left caudal ventrolateral precentral gyrus (Caudal ventrolateral PrG, 177 voxels), caudal cuneus (Caudal Cun, 12600 voxels), left superior temporal gyrus (Planum) (STG(Planum), 500 voxels), left superior temporal gyrus (anterior division) (STG (anterior division), 137 voxels), paracentral lobule (PCL, 110 voxels). **b**. Clusters with significant between-group differences in the drug cue-reactivity contrast between MUDs and HCs. All clusters survived the FDR-corrected p<0.05 threshold, and have higher drug>neutral activations in MUDs compared to HCs (indicated by warm colors). Four significant clusters were found: Medial orbital gyrus (Medial OrG, 323 voxels), left inferior frontal junction (IFJ, 307 voxels), left lateroventral fusiform gyrus (Lateroventral FuG, 179 voxels), right lateral occipital cortex (LOcC, 160 voxels) **c**. The bar chart in the clusters with significant differences between MUDs and HCs in drug cue-reactivity contrast. The bars estimate the mean z-value in the drug cue-reactivity contrast in each cluster. The error bars show the standard error of z-statistic values across 53 MUDs and 23 HCs. HCs: healthy controls, MUDs, methamphetamine use disorders.

To test the validation of the task, we compared brain activations of drug cue-reactivity (*Drug>Neutral*) contrast between MUDs and HCs. MUDs had a higher drug cue-reactivity than HCs in the left medial orbital gyrus (323 voxels, z-max=4.19), left inferior frontal junction (307 voxels, z-max=4.64), left lateroventral fusiform gyrus (179 voxels, z-max=4.12) and the right lateral occipital cortex (160 voxels, z-max=4.09), corrected p-value<0.05 (Figure 3.b and 3.c and Table S3).

Negative versus neutral cues were associated with higher activations in the cuneus (16159 voxels, z-max=8.92), bilateral ORG (left: 229 voxels, z-max=4.85), (right: 291 voxels, z-max=5.67) left Inferior Frontal Gyrus (IFG) (259 voxels, z-max=4.42), bilateral medial amygdala (left:163 voxels, z-max=5.67), (right=189 voxels, z-max=6.23), left occipital thalamus (185 voxels, z-max=4.56), and left lateral SFG (182 voxels, z-max=5.36), and lower activations in the left postcentral gyrus (POG) (5059 voxels, z-max=5.69), right caudal IPL (2931 voxels, z-max=6.11), ventromedial middle frontal gyrus (MFG) (2105 voxels, z-max=5.04), and bilateral bilateral intermediate lateral ITG (left: 318 voxels, z-max=5.08, right: 1032 voxels, z-max=5.45), dorsal CG (907 voxels, z-max=5.24), Operculum IFG (635 voxels, z-max=4.99), with a corrected p-value<0.05 (Figure 4, and Table S4).

**Figure 4:**
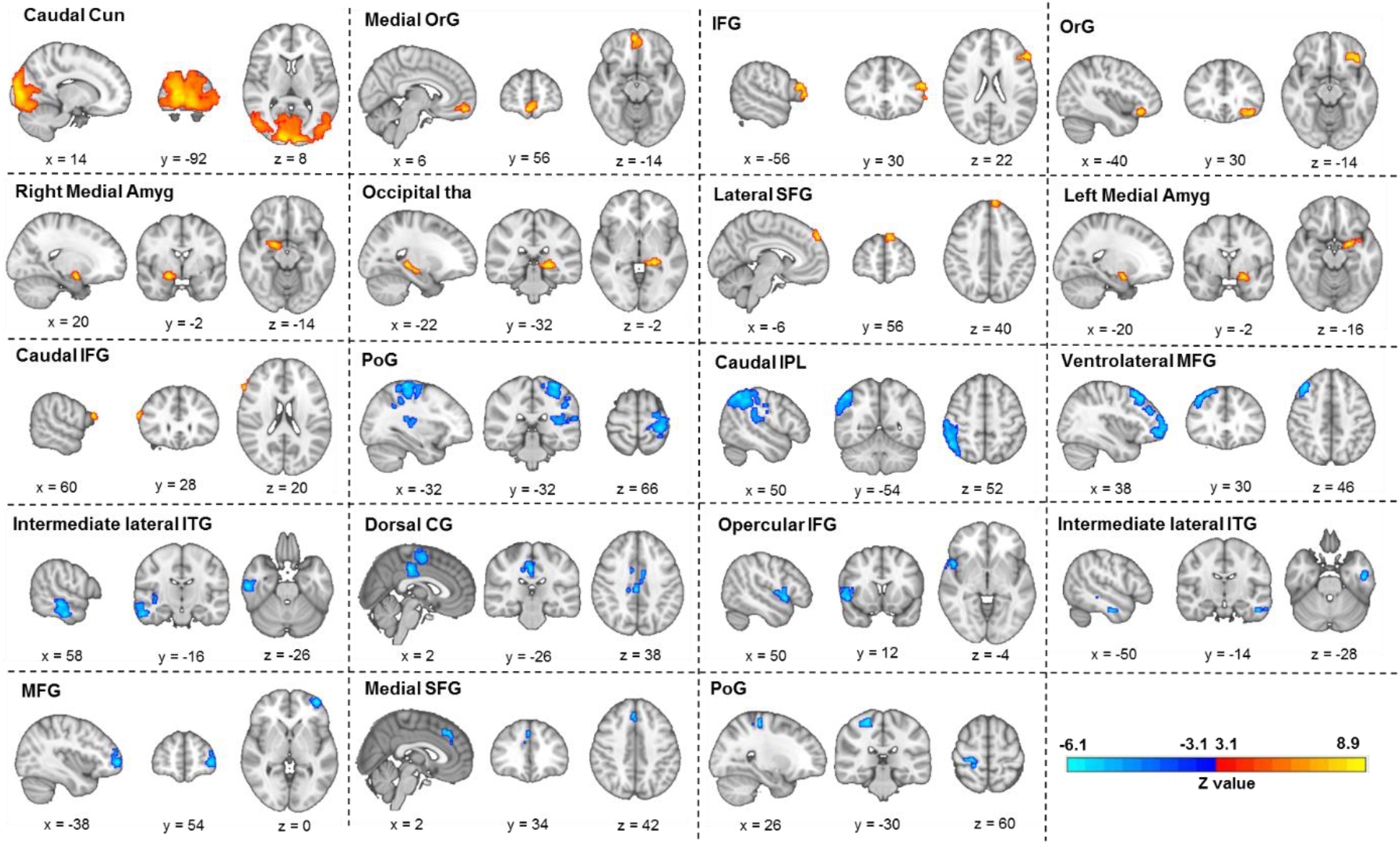
Negative cue reactivity. Significant clusters in the negative cue-reactivity whole-brain contrast in MUDs surviving the FDR-corrected p<0.05 threshold. Voxels with positive z-values are displayed with warm colors (red to yellow) and show a higher activation associated with negative compared to neutral cues, while voxels with negative z-values are displayed with cold colors (blue to cyan) and have a higher activation when viewing neutral compared to negative cues. MNI coordinate of the peak voxel in each significant cluster is reported bellow sagittal, coronal, and axial views of the brain. 19 significant clusters are visualized based on cluster-defining threshold method and reported based on Brainnetome atlas parcellation as follow: caudal cuneus (Caudal Cun, 16159 voxels), medial orbital gyrus (Medial OrG, 291 voxels), left inferior frontal gyrus (IFG, 259 voxels), left orbital gyrus (OrG, 229 voxels), medial amygdala (Right Medial Amyg, 189 voxels, Left Medial Amyg, 163 voxels), left occipital thalamus (Occipital tha, 185 voxels), left lateral superior frontal gyrus (Lateral SFG, 182 voxels), right caudal inferior frontal gyrus (Caudal IFG, 128 voxels), left postcentral gyrus (PoG, 5059 voxels), right caudal inferior parietal lobule (Caudal IPL, 2931 voxels), right ventrolateral middle frontal gyrus (Ventrolateral MFG, 2105 voxels), intermediate lateral inferior temporal gyrus (Right Intermediate lateral ITG, 1032 voxels, left Intermediate lateral ITG, 318 voxels), Dorsal Cingulate Gyrus (Dorsal CG, 907 voxels), right Operculum (Opercular IFG, 635 voxels), left middle frontal gyrus (MFG, 195 voxels), right medial superior frontal gyrus (Medial SFG, 157 voxels), right postcentral gyrus (PoG, 156 voxels).

### 3.4 Response-inhibition

In the overall No-Go>Go response inhibition contrast *(DSNG+NSNG+BSNG+NESNG) > (DSG+NSG+BSG+NESG)*, activations survived a corrected p-value<0.05 threshold in the left rostroventral IPL (1522 voxels, z-max=5.64), right anterior superior MTG (1515 voxels, z-max=5.36), left medial precuneus (Pcun) (777 voxels, z-max=4.57), left lateral SFG (169 voxels, z-max=4.34), right rostral IFG (124 voxels, z-max=4.38) and right pregenual CG (116 voxels, z-max=4.09). Go trials were associated with higher acrivations than No-Go trials in the right medioventral FUG (3305 voxels, z-max=6.7), Left POG (2632 voxels, z-max=6.87), and Left caudal cuneus (624 voxels, z-max=5.97), right medial SFG (381 voxels, z-max=5.48), left Caudal ventrolateral PrG (346 voxels, z-max=4.9), left Ventral Caudate (322 voxels, z-max=4.6), left Medioventral Fusiform Gyrus FuG (297 voxels, z-max=4.95), left Inferior DLPFC (95 voxels, z-max=3.67), (Figure 5, and Table S5).

**Figure 5:**
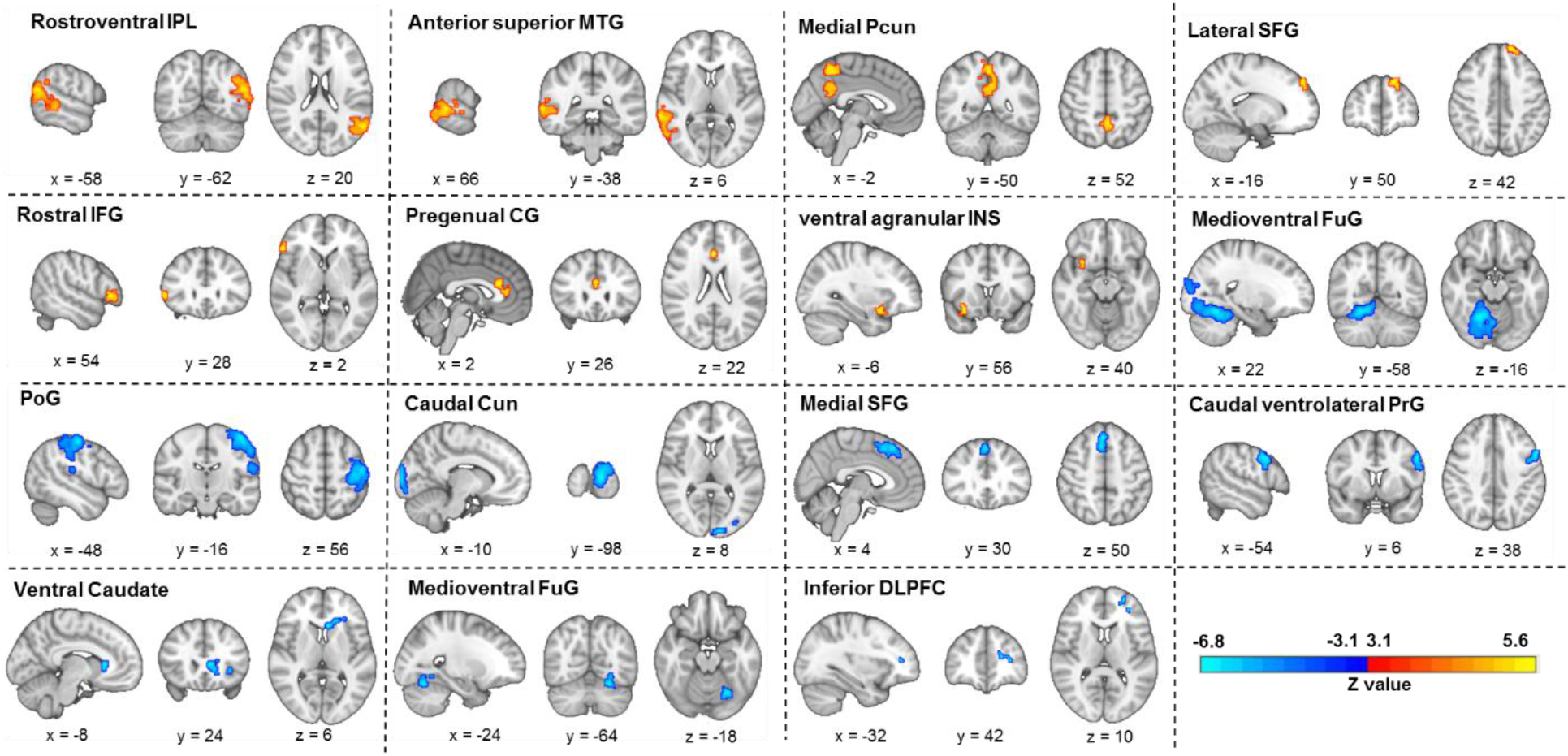
Response inhibition. Significant clusters in the response-inhibition whole-brain contrast in MUDs surviving the FDR-corrected p<0.05 threshold. Voxels with positive z-values are displayed with warm colors (red to yellow) and show a higher activation associated with No-Go compared to Go trials, while voxels with negative z-values are displayed with cold colors (blue to cyan) and have a higher activation during Go compared to No-Go trials. MNI coordinate of the peak voxel in each significant cluster is reported bellow sagittal, coronal, and axial views of the brain. 15 significant clusters are visualized based on cluster-defining threshold method and reported based on Brainnetome atlas parcellation as follow: left rostroventral inferior parietal lobule (Rostroventral IPL, 1552 voxels), right anterior superior middle temporal gyrus (Anterior superior MTG, 1515 voxels), middle precuneus (Middle Pcun, 777 voxels), left lateral superior frontal gyrus (Lateral SFG, 169 voxels), right rostral inferior frontal gyrus (Rostral IFG, 124), pregenual cingulate gyrus (Pregenual CG, 116 voxels), right ventral agranular insular gyrus (Ventral agranular INS, 71 voxels), right medioventral fusiform gyrus (Medioventral FuG, 3305 voxels), left postcentral gyrus (PoG, 2632 voxels), left caudal cuneus (Caudal Cun, 624 voxels), medial superior frontal gurus (Medial SFG, 381 voxels), left caudal ventrolateral precentral gyrus (Caudal ventrolateral PrG, 346 voxels), left ventral caudate (Ventral Caudate, 322 voxels), left medioventral fusiform gyrus (Medioventral FuG, 297 voxels), left inferior dorsolateral prefrontal cortex (inferior DLPFC, 95 voxels).

### 3.5 Response-inhibition during drug cue-reactivity and negative emotional cue-reactivity

The drug cue-reactivity/response-inhibition interaction contrast revealed a higher activation during drug-related inhibition than neutral cue inhibition in the left caudal cuneus (5129 voxels, z-max=6.94), right dorsomedial parietooccipital Pcun (137 voxels, z-max=4.28), right medial superior occipital gyrus (116 voxels, z-max=4.05) and left LOcC (98 voxels, z-max=3.67), and with lower activation in the bilateral POG (left: 1647 voxels, z-max=4.49, right: 319 voxels, z-max=4.54), right caudodorsal CG (1317 voxels, z-max=4.61), right rostroventral IPL (776 voxels, z-max=4.36), left medial SFG (352 voxels, z-max=4.15), and right dorsal agranular insula (152 voxels, z-max=4.32), corrected p-value<0.05 (Figure 6, and Table S6).

**Figure 6:**
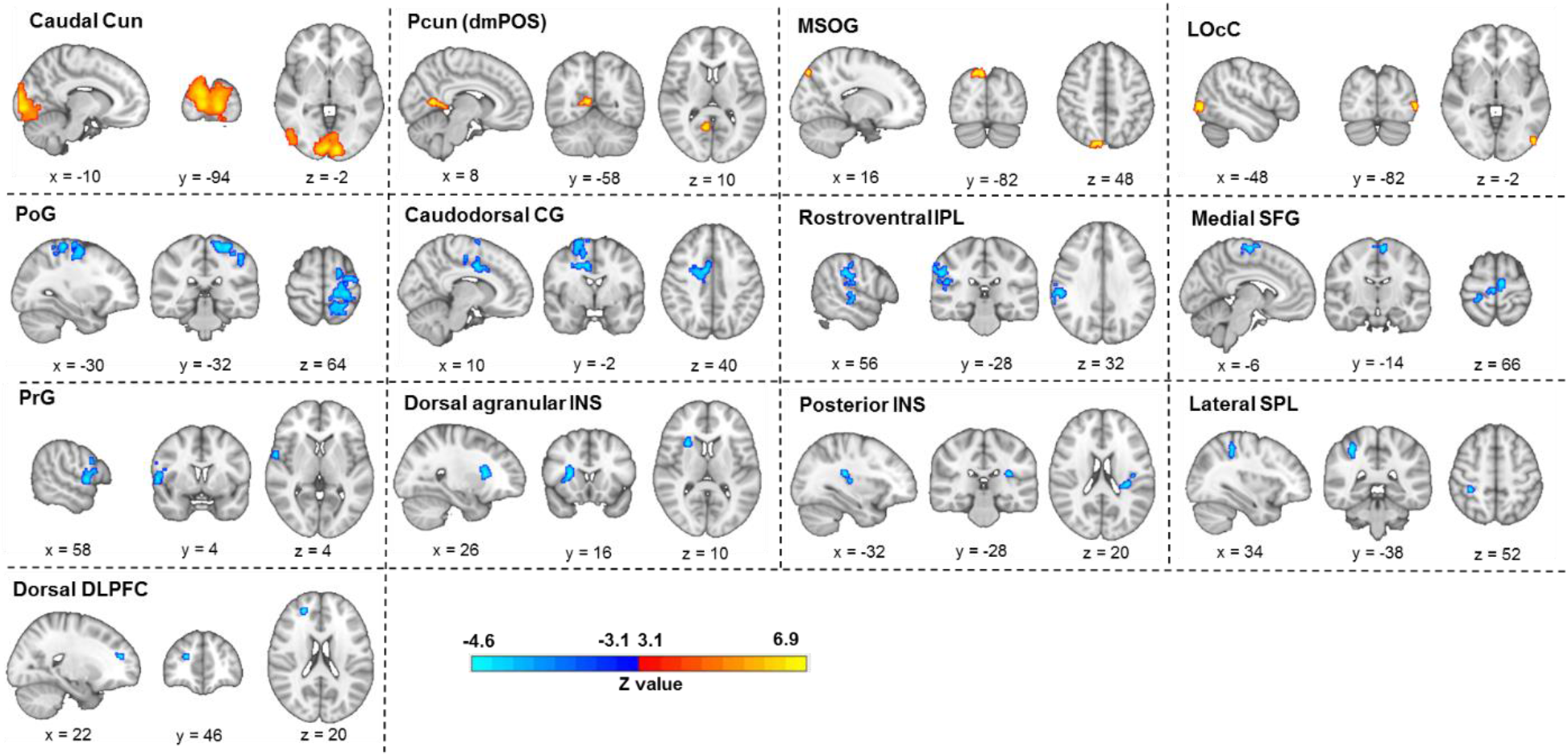
Drug cue reactivity/ response inhibition interaction. Significant clusters in the drug cue-reactivity/response-inhibition interaction whole-brain contrast in MUDs surviving the FDR-corrected p<0.05 threshold. Voxels with positive z-values are displayed with warm colors (red to yellow) and show a higher activation associated with response-inhibition during exposure to methamphetamine compared to neutral cues, while voxels with negative z-values are displayed with cold colors (blue to cyan) and have a higher activation during response-inhibition when viewing neutral compared to methamphetamine cues. MNI coordinate of the peak voxel in each significant cluster is reported bellow sagittal, coronal, and axial views of the brain. 13 significant clusters are visualized based on cluster-defining threshold method and reported based on Brainnetome atlas parcellation as follow: caudal cuneus (Caudal Cun, 5129 voxels), right dorsomedial parietooccipital precuneus (Pcun (dmPOS), 137 voxels), right medial superior occipital gyrus (MSOG, 116 voxels), left lateral occipital cortex (LOcC, 98 voxels), left postcentral gyrus (PoG, 1647 voxels), right caudodorsal cingulate gyrus (Caudodorsal CG, 1317 voxels), right rostroventral inferior parietal lobule (Rostroventral IPL, 776 voxels), medial superior frontal gyrus (Medial SFG, 352 voxels), right precentral gyrus (PrG, 319 voxels), right dorsal agranular insula (Dorsal agranular INS, 152 voxels), left posterior insular gyrus (Posterior INS, 124 voxels), left lateral superior parietal lobule (Lateral SPL, 84 voxels), left dorsal dorsolateral prefrontal cortex (Dorsal DPLFC, 69 voxels).

Response-inhibition during negative cue exposure is associated with greater activity compared to neutral inhibition in the right dorsolateral MTG (369 voxels, z-max=4.27), and left rostroventral IPL (125 voxels, z-max=4.11), and lower activity only in the left Lateroventral Fusiform Gyrus (277 voxels, z-max=4.43), corrected p-value<0.05 (Figure 7 and Table S7).

**Figure 7:**
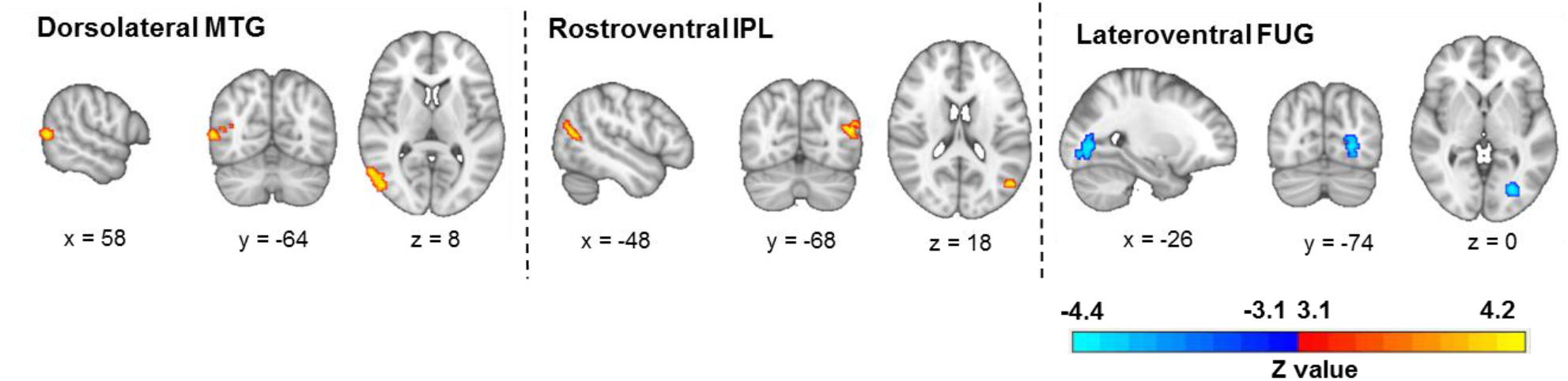
Negative cue reactivity/ response inhibition interaction. Significant clusters in the negative cue-reactivity/response-inhibition interaction whole-brain contrast in MUDs surviving the FDR-corrected p<0.05 threshold. Voxels with positive z-values are displayed with warm colors (red to yellow) and show a higher activation associated with response-inhibition during exposure to negative compared to neutral cues, while voxels with negative z-values are displayed with cold colors (blue to cyan) and have a higher activation during response-inhibition when viewing neutral compared to negative cues. MNI coordinate of the peak voxel in each significant cluster is reported bellow sagittal, coronal, and axial views of the brain. 3 significant clusters are visualized based on cluster-defining threshold method and reported based on Brainnetome atlas parcellation as follow: right dorsolateral middle temporal gyrus (Dorsolateral MTG, 369 voxels), left rostroventral inferior parietal lobule (Rostroventral IPL, 125 voxels) and left lateroventral fusiform gyrus (Lateroventral FUG, 277 voxels)

### 3.6 Correlates of brain activation

The negative interaction contrast in the right dorsolateral MTG cluster (beta=-0.357, p-corrected=0.018) had Bonferroni-corrected significant correlations with the pre-scanning negative PANAS and BIS motor inhibition scores (beta=-0.319, p-corrected=0.04). No correlations between the cue-reactivity contrast and behavioral or clinical variables survived FDR correction, but there are uncorrected positive correlations between cue-reactivity and pre-scanning craving self-report (VAS) in the left lateral SFG and left dorsal CG (beta=0.28, p-uncorrected=0.036), with post-scanning VAS in the left ORG (beta=0.303, p-uncorrected=0.027), with commission error rate in the left dorsal CG (beta=0.307, p-uncorrected=0.025), with omission error rate in the left caudal ventrolateral PRG (beta=0.295 p-uncorrected=0.032) and the left STG (beta=0.277 p-uncorrected=0.045), and uncorrected negative correlations in right caudal cuneus (beta=-0.272, p-uncorrected=0.049). Response-inhibition during drug cue reactivity has an uncorrected negative correlation with commission error in the left caudal cuneus (beta=-0.301, p-uncorrected=0.028), and positive correlation in the dorsal MFG (beta=0.282, p-uncorrected=0.041). Response-inhibition has uncorrected negative correlations with meth use duration in the left medioventral FUG (beta=-0.28, p-uncorrected=0.036), and with commission error in the Left medial Precuneus (beta=-0.291, p-uncorrected=0.034), as well as negative correlations with omission error in the left medioventral FUG (beta=-0.316, p-uncorrected=0.021) and left caudal cuneus (beta=-0.369, p-uncorrected=0.006) (Table S8).

## 4. Discussion

This study adds to the scant literature on the interaction of response-inhibition and reactivity to negative and drug cues in individuals with MUD, developing the first mixed fMRI response inhibition task which can assess response inhibition in the presence of negative and drug cue reactivity in individuals with methamphetamine use disorder (MUD). Subsequently, we validated this task in a sample of participants with MUD by demonstrating behavioural and neural evidence of the influence of negative and drug cue presentation on response inhibition. The implications of the observed brain activations and deactivations will be discussed below, along with their correlations with subjective craving and markers of inhibitory performance.

### 4.1 Cue-reactivity

Drug cue-reactivity was associated with activations in the striatum and the superior frontal and pregenual anterior cingulate gyri (CG). While the striatum and anterior CG are important nodes in the reward network (Luijten et al., 2017; Zilverstand et al., 2018), the SFG is broadly implicated in decision making (Boisgueheneuc et al., 2006). The observed SFG activation may be related to reward-related decision making during drug cue-reactivity, as SFG activity is influenced by dopaminergic pathways during reward appraisal (Ott and Nieder, 2019) and has been observed in SUDs (Garavan et al., 2000; Grüsser et al., 2004). The IPL and dorsal CG had increased activations during drug cue-reactivity but decreased activity in negative emotional cue-reactivity. As nodes of the salience network (Zilverstand et al., 2018), the IPL and dorsal CG are involved in drug craving and drug-seeking (Kühn and Gallinat, 2011; Naqvi and Bechara, 2009), and the different effects of drug cue-reactivity and negative emotional cue-reactivity on their activity may indicate the redirecting of attentional resources towards drug cues and away from negative cues (Corbetta et al., 2008; Shenhav et al., 2013). On the other hand, the IFG was activated in both drug cue-reactivity and negative emotional cue-reactivity. The IFG’s involvement in drug cue-reactivity has been reported before (Hanlon et al., 2018; Reuter et al., 2005), but it also plays a vital role in processing conflicts between opposing representations and may have been engaged by our complicated task design to resolve the resulting conflicts (Novick et al., 2005; Swick et al., 2008). Notably, the MUDs also differed from HCs in the drug cue-reactivity-associated activation of the inferior frontal junction, which borders the IFG.

Lastly, we also observed altered drug cue-reactivity and negative cue-reactivity-associated activations in the cuneus, which is part of the default mode network (DMN) and is involved in episodic memory retrieval (Xu et al., 2016), and the amygdala, central to the processing of salient stimuli as part of the limbic network (Chase et al., 2011; Dickerson and Eichenbaum, 2010; Tang et al., 2012). Increased functional activity in the posterior part of the DMN, together with decreased MFG activation, suggests the engagement of self-referential processing during negative emotional cue-reactivity (Xu et al., 2016). Increased amygdalar and cuneal activation in response to negative versus neutral cues was also observed in HCs, suggesting that negative emotional cue-reactivity can generally engage emotional and self-referential processing in all participants.

### 4.2 Response-inhibition

Response-inhibition was associated with activations in the left IPL and SFG, and right MTG, IFG and the anterior cingulate cortex. All these regions are known to be involved in successful response-inhibition, supporting the validity of the task and the chosen contrast (Aron, 2007; Dong et al., 2012; Duann et al., 2009; Gruber and Yurgelun-Todd, 2005; Hampshire et al., 2010; Mostofsky and Simmonds, 2008; Tapert et al., 2007). Notably, the right-lateralization of IFG and IPL, import nodes in the inhibitory frontoparietal network, has been observed in previous research on response-inhibition as well (Garavan et al., 1999; Garavan et al., 2002; Hampshire et al., 2010; McNab et al., 2008). We also observed occipital activations which, while more rarely observed, have been reported in some previous studies (Braver et al., 2001; Kelly et al., 2004; Liddle et al., 2001; Mathalon et al., 2003; Wager et al., 2005). Despite these similarities, caution should be exercised when comparing the neural substrates of response-inhibition across studies, as the fMRI activation patterns associated with response-inhibition are task-dependent (Swick et al., 2011; Yeung et al., 2020), and different response-inhibition tasks may variably engage associated processes such as response selection (Simmonds et al., 2008). For example, while most of the cited research suggests a hyperactivation of inhibitory regions in SUDs, some studies suggest reverse alterations in response-inhibition-associated neural activity in SUDs (Wallace et al., 2020).

### 4.3 Interaction of response-inhibition, drug cue-reactivity and negative emotional cue-reactivity

Higher commission and omission error rates and longer reaction times in drug and negative cue blocks indicate an impairing influence of drug cue-reactivity and negative emotional cue-reactivity on response-inhibition. Such inhibitory impairments have been observed frequently on studies of response-inhibition during exposure to drug (Czapla et al., 2015; Lannoy et al., 2019; Noël et al., 2007; Weafer and Fillmore, 2012) or emotionally negative cues (Albert et al., 2010; Goldstein et al., 2007; Ramos-Loyo et al., 2017). It has been suggested that response-inhibition may require more resources when processing negative valence (Goldstein et al., 2007), and in the case of drug cues, this interaction might be partly mediated by approach bias (Kreusch et al., 2013) or increased cognitive impulsivity in the presence of drugs (Noël et al., 2007).

Response-inhibition during drug cue-reactivity was associated with higher activations in the bilateral visual cortices and cunei and lower activations in regions associated with response-inhibition (MFG and IPL), motor control (precentral gyrus, supplementary motor area) and interoception (insula). Some previous studies on the interaction of response inhibition and reactivity to appetitive cues suggest that reduced activation of inhibitory regions may indicate poorer recruitment of inhibitory control resources during cue-reactivity (Batterink et al., 2010; Gilman et al., 2018; Liu et al., 2014; Seok and Sohn, 2020). Higher activations across the occipital cortices have also been observed in mixed Go/No-Go cue-reactivity studies on alcohol-dependent individuals, potentially since the higher salience and complexity of substance No-Go cues necessitate more visual processing (Czapla et al., 2017; Stein et al., 2021). It is notable that MUDs also had a higher drug cue-reactivity-related occipital activation than HCs, supporting the relevance of these activations to the disorder.

Response-inhibition was associated with a higher activation when viewing negative compared to neutral cues in the right MTG and left IPL. Dysfunctional activation patterns in the MTG and IPL, especially the supramarginal gyrus, have been implicated in response-inhibition impairments in individuals with addictive disorders (Chikazoe et al., 2007; Qiu and Wang, 2021) and response-inhibition when viewing negative cues (Brown et al., 2012; Chester et al., 2016; Goldstein et al., 2007). Notably, right MTG activation was also significantly correlated with pre-scanning negative affect and self-reported motor impulsivity, indicating that the MTG may be an essential hub for the interaction of negative affective processing and response-inhibition. More investigations of the role of IPL and MTG activity in negative urgency in MUD are warranted given the broader involvement of these regions in the neurobiology of the disorder (Jan et al., 2012; Paulus and Stewart, 2020; Sabrini et al., 2019).

### 4.4 Limitations

This project has a number of notable limitations. In terms of participants, the sample size is substantial but includes no female subjects, and while data from HCs was used only to validate the reactivity of MUD participants to methamphetamine cues, sample sizes were unbalanced. The MUD participants were also heterogeneous in certain respects, and the methamphetamine use duration had a high variance. Regarding the task design, factorial and mixed tasks limit the statistical power of inference on different conditions of interest.

### 4.5 Conclusion

Besides adding to the literature on the neural substrates of response-inhibition and reactivity to emotionally negative and drug-associated cues in MUD, this is the first study on the fMRI-correlates of the interactions of these phenomena in this addictive disorder. There is already some evidence that drug cue-reactivity and response-inhibition fMRI paradigms may help develop biomarkers to aid diagnosis, prognosis, and treatment monitoring (Bough et al., 2014; Garrison and Potenza, 2014; Zilverstand et al., 2018). Studies using mixed response-inhibition tasks may further aid translational efforts (Noël et al., 2007), and recent research suggests that response-inhibition during cue reactivity can elicit neural activations that predict clinically relevant outcomes such as relaps (Gilman et al., 2018). Considering the engagement of various regions known to be involved in the neurobiology of SUDs by our mixed Go/No-Go task and the modulation of response-inhibitory activity by cue-induced craving and negative affect, further consideration of these paradigms is warranted. Future research should use longitudinal designs and multicentric scanning to assess the reliability and generalisability of various activation patterns, measure clinical outcomes and their co-variation with these patterns, and use these patterns, alone or in combination with parameter estimates derived from other paradigms, to develop and test biomarkers with potential for clinical translation.

## Data Availability

Anonymized data for this manuscript is available upon request from the corresponding author.

## Supplementary Materials

**Table S1.** Behavioral performance

| Behavioral Measure | Task-Block (Mean (SD)) |  |  |  | ANOVA P-value |
| --- | --- | --- | --- | --- | --- |
|  | Blank | Neutral | Negative | Drug |  |
| Commission Error Percentage | 6.53 (7.9) | 11.32 (10.41) | 12.50 (11.19) | 13.29 (13.76) | 0.009 |
| Omission Error Percentage | 3.09 (3.75) | 9.93 (10.50) | 12.42 (10.16) | 12.92 (10.99) | 0.0001 |
| Reaction Time (sec) | 0.67 (0.07) | 0.76 (0.06) | 0.77 (0.05) | 0.76 (0.06) | 0.0001 |

**Note.** Values are expressed as “averages (standard error of the mean)”. The p-value of the single-factor ANOVA test represents significant differences. Omission error is defined as failing to respond to Go trials, and commission error is the failure to inhibit pre-potent responses on No-Go trials.

**Table S2:**
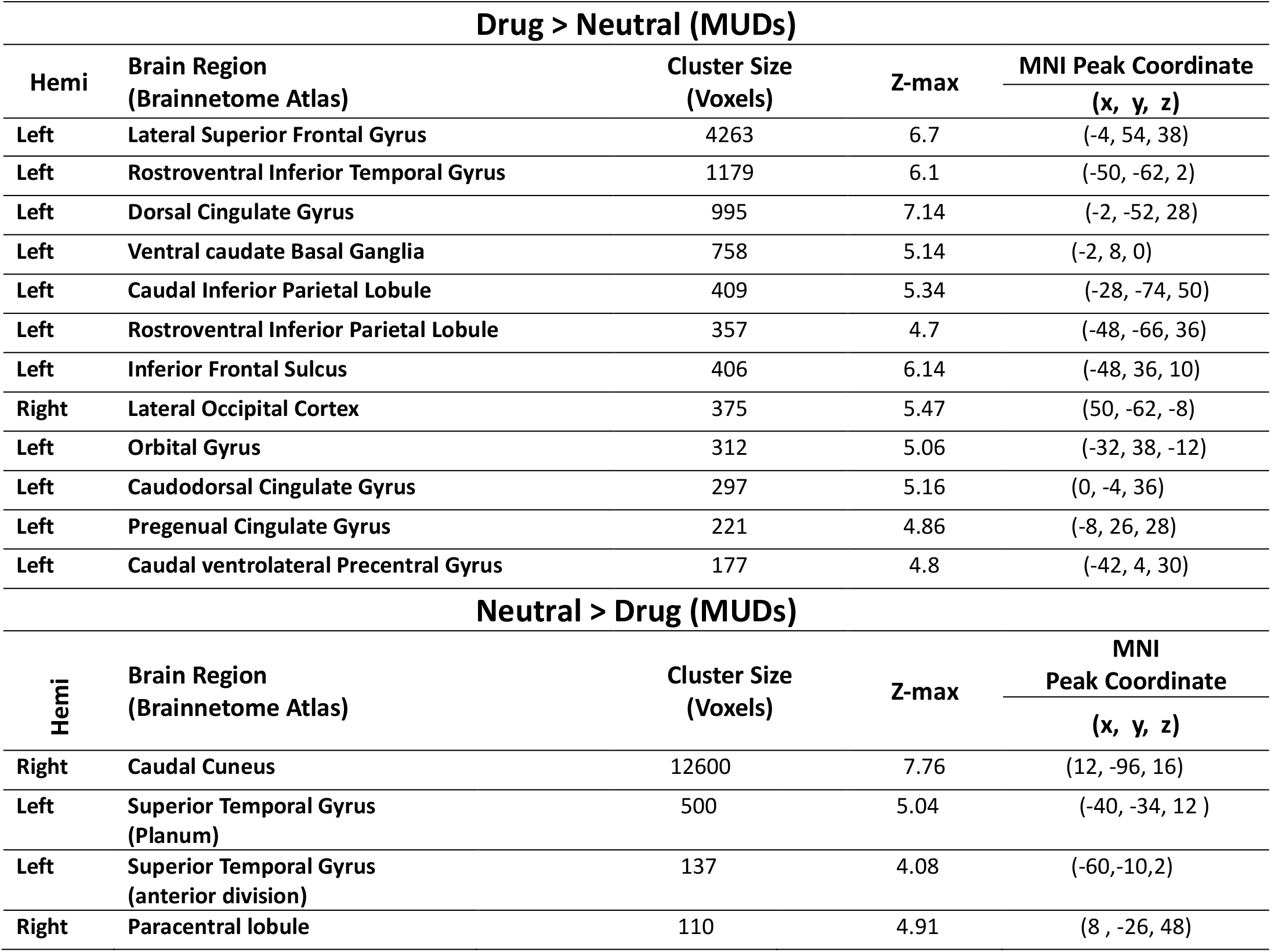
Significant clusters in the drug cue-reactivity whole-brain contrast in MUDs based on the Cluster Defining Threshold method (corrected p-value <0.05). Cluster size, z-max values, and MNI coordinates for voxels with maximal z-values in each cluster are presented. Clusters with a higher activation associated with methamphetamine compared to neutral cues and those with higher activation when viewing neutral compared to drug cues are listed separately. Hemi: hemisphere.

**Table S3:** Clusters with significant between-group differences in the drug cue-reactivity contrast between MUDs and HCs, based on Cluster Defining Threshold method (corrected p-value <0.05). Cluster size, z-max values, and MNI coordinates for voxels with maximal z-values in each cluster are presented. All clusters have higher drug>neutral activations in MUDs compared to HCs. Hemi: hemisphere.

| Group Difference Drug Cue-Reactivity (MUD > HC) |  |  |  |  |
| --- | --- | --- | --- | --- |
| Hemi | Brain Region<br>(Brainnetome Atlas) | Cluster Size<br>(Voxels) | Z-max | MNI |
|  |  |  |  | Peak Coordinate<br>(x, y, z) |
| Left | Medial Orbital Gyrus | 323 | 4.19 | (-4, -56, -0) |
| Left | Inferior Frontal Junction | 307 | 4.64 | (-36, 2, 30) |
| Left | Lateroventral Fusiform Gyrus | 179 | 4.12 | (-44, -68, -10) |
| Right | Lateral Occipital Cortex | 160 | 4.09 | (58, -72, -8) |

**Table S4:** Significant clusters in the negative cue-reactivity whole-brain contrast in MUDs based on the Cluster Defining Threshold method (corrected p-value <0.05). Cluster size, z-max values, and MNI coordinates for voxels with maximal z-values in each cluster are presented. Clusters with a higher activation associated with negative emotional compared to neutral cues and those with higher activation when viewing neutral compared to negative emotional cues are listed separately.

| Negative > Neutral (MUDs) |  |  |  |  |
| --- | --- | --- | --- | --- |
| Hemi | Brain Region<br>(Brainnetome Atlas) | Cluster Size<br>(Voxels) | Z-max | MNI |
|  |  |  |  | Peak Coordinate<br>(x, y, z) |
| Right | Caudal Cuneus | 16159 | 8.92 | (14, -92, 8) |
| Right | Medial Orbital Gyrus | 291 | 5.67 | (6, 56, -14) |
| Left | Inferior Frontal Gyrus | 259 | 4.42 | (-56, 30, 22) |
| Left | Orbital Gyrus | 229 | 4.85 | (-40, 30, -14) |
| Right | Medial Amygdala | 189 | 6.23 | (20, -2, -14) |
| Left | Occipital Thalamus | 185 | 4.56 | (-22, -32, -2) |
| Left | Lateral Superior Frontal Gyrus | 182 | 5.36 | (-6, 56, 40) |
| Left | Medial Amygdala | 163 | 5.67 | (-20, -2, -16) |
| Right | Caudal Inferior Frontal Gyrus | 128 | 4.71 | (60, 28, 20) |
| Neutral > Negative (MUDs) |  |  |  |  |
| Left | Postcentral Gyrus | 5059 | 5.69 | (-32, -32, 66) |
| Right | Caudal Inferior Parietal Lobule | 2931 | 6.11 | (50, -54, 52) |
| Right | Ventrolateral Middle Frontal Gyrus | 2105 | 5.04 | (38, 30, 46) |
| Right | Intermediate lateral Inferior Temporal Gyrus | 1032 | 5.45 | (58, -16, -26) |
| Right | Dorsal Cingulate Gyrus | 907 | 5.24 | (2, -26, 38) |
| Right | Operculum IFG | 635 | 4.99 | (50, 12, -4) |
| Left | Intermediate lateral Inferior Temporal Gyrus | 318 | 5.07 | (-50, -14, -28) |
| Left | Middle Frontal Gyrus | 195 | 5 | (-38, 54, 0) |
| Right | Medial Superior Frontal Gyrus | 157 | 4.2 | (2, 34, 42) |
| Right | Postcentral Gyrus | 156 | 4.18 | (26, -30, 60) |

**Table S5:** Significant clusters in the response-inhibition whole-brain contrast in MUDs based on the Cluster Defining Threshold method (corrected p-value <0.05). Cluster size, z-max values, and MNI coordinates for voxels with maximal z-values in each cluster are presented. Clusters with a higher activation associated with No-Go trials compared to Go trials and those with higher activation during Go compared to No-Go trials are listed separately. Hemi: hemisphere.

| **Successful No-Go (Blank + Neutral + Negative + Drug) > Successful Go (Blank + Neutral + Negative + Drug)** |  |  |  |  |
| --- | --- | --- | --- | --- |
| **Hemi** | **Brain Region**<br>**(Brainnetome Atlas)** | **Cluster Size**<br>**(Voxels)** | **Z-max** | **MNI** |
|  |  |  |  | **Peak Coordinate**<br>**(x, y, z)** |
| Left | Rostroventral Inferior Parietal Lobule | 1552 | 5.64 | (-58, -62, 20) |
| Right | Anterior Superior Middle Temporal Gyrus | 1515 | 5.36 | (66, -38, 6) |
| Left | Medial Precuneus | 777 | 4.57 | (-2, -50, 52) |
| Left | Lateral Superior Frontal Gyrus | 169 | 4.34 | (-16, 50, 42) |
| Right | Rostral Inferior Frontal Gyrus | 124 | 4.38 | (54, 28, 2) |
| Right | Pregenual Cingulate Gyrus | 116 | 4.09 | (2, 26, 22) |
| Right | Ventral agranular Insular Gyrus | 71 | 3.9 | (32, 14, -14) |
| **Successful Go (Blank + Neutral + Negative + Drug) > Successful No-Go (Blank + Neutral + Negative + Drug)** |  |  |  |  |
| Right | Medioventral Fusiform Gyrus | 3305 | 6.7 | (22, -58, -16) |
| Left | Postcentral Gyrus | 2632 | 6.87 | (-48, -16, 56) |
| Left | Caudal Cuneus | 624 | 5.97 | (-10, -98, 8) |
| Right | Medial Superior Frontal Gyrus | 381 | 5.48 | (4, 30, 50) |
| Left | Caudal Ventrolateral Precentral Gyrus | 346 | 4.9 | (-54, 6, 38) |
| Left | Ventral Caudate | 322 | 4.6 | (-8, 24, 6) |
| Left | Medioventral Fusiform Gyrus | 297 | 4.95 | (-24, -64, -18) |
| Left | Inferior DLPFC | 95 | 3.67 | (-32, 42, 10) |

**Table S6:** Significant clusters in the drug cue-reactivity/response-inhibition interaction whole-brain contrast in MUDs based on the Cluster Defining Threshold method (corrected p-value <0.05). Cluster size, z-max values, and MNI coordinates for voxels with maximal z-values in each cluster are presented. Clusters with a higher activation associated with response-inhibition during exposure to methamphetamine compared to neutral cues and those with higher activation during response-inhibition when viewing neutral compared to methamphetamine cues are listed separately. Hemi: hemisphere.

| **(Successful No-Go Drug > Successful Go Drug) > (Successful No-Go Neutral > Successful Go Neutral)** |  |  |  |  |
| --- | --- | --- | --- | --- |
| **Hemi** | **Brain Region**<br>**(Brainnetome Atlas)** | **Cluster Size**<br>**(Voxels)** | **Z-max** | **MNI Peak Coordinate** |
|  |  |  |  | **(x, y, z)** |
| Left | Caudal Cuneus | 5129 | 6.94 | (-10, -94, -2) |
| Right | Dorsomedial parietooccipital Precuneus | 137 | 4.28 | (8, -58, 10) |
| Right | Medial Superior Occipital Gyrus | 116 | 4.05 | (16, -82, 48) |
| Left | Lateral Occipital Cortex | 98 | 3.67 | (-48, -82, -2) |
| **(Successful No-Go Neutral > Successful Go Neutral) > (Successful No-Go Drug > Successful Go Drug)** |  |  |  |  |
| Left | Postcentral Gyrus | 1647 | 4.49 | (-30, -32, 64) |
| Right | Caudodorsal Cingulate Gyrus | 1317 | 4.61 | (10, -2, 40) |
| Right | Rostroventral Inferior Parietal Lobule | 776 | 4.36 | (56, -28, 32) |
| Left | Medial Superior Frontal Gyrus | 352 | 4.15 | (-6, -14, 66) |
| Right | Precentral Gyrus | 319 | 4.54 | (58, 4, 4) |
| Right | Dorsal agranular insula | 152 | 4.32 | (26, 16, 10) |
| Left | Posterior Insular Gyrus | 124 | 4.09 | (-32, -28, 20) |
| Right | Lateral Superior Parietal Lobule | 84 | 3.91 | (34, -38, 52) |
| Right | Dorsal DLPFC | 69 | 3.67 | (22, 46, 20) |

**Table S7:**
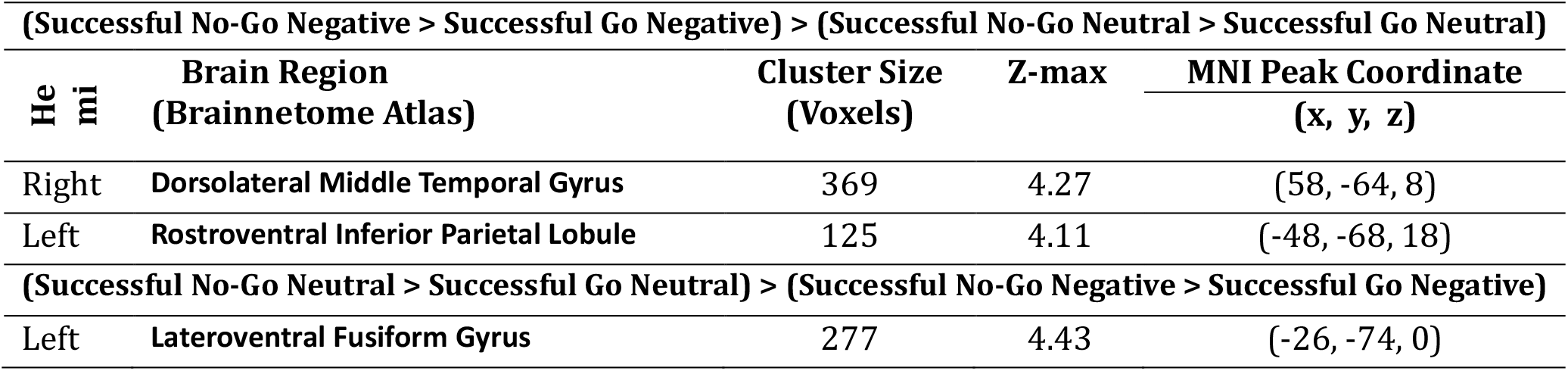
Significant clusters of the interaction between Negative Cue-Reactivity and Response Inhibition in MUDs based on Cluster Defining Threshold method (corrected p-value <0.05, *: survived with p-value <.001). Hemi: hemisphere.

| **(Successful No-Go Negative > Successful Go Negative) > (Successful No-Go Neutral > Successful Go Neutral)** |  |  |  |  |
| --- | --- | --- | --- | --- |
| Hemi | Brain Region<br>(Brainnetome Atlas) | Cluster Size<br>(Voxels) | Z-max | MNI Peak Coordinate<br>(x, y, z) |
| Right | Dorsolateral Middle Temporal Gyrus | 369 | 4.27 | (58, -64, 8) |
| Left | Rostroventral Inferior Parietal Lobule | 125 | 4.11 | (-48, -68, 18) |
| **(Successful No-Go Neutral > Successful Go Neutral) > (Successful No-Go Negative > Successful Go Negative)** |  |  |  |  |
| Left | Lateroventral Fusiform Gyrus | 277 | 4.43 | (-26, -74, 0) |

**Table S8:**
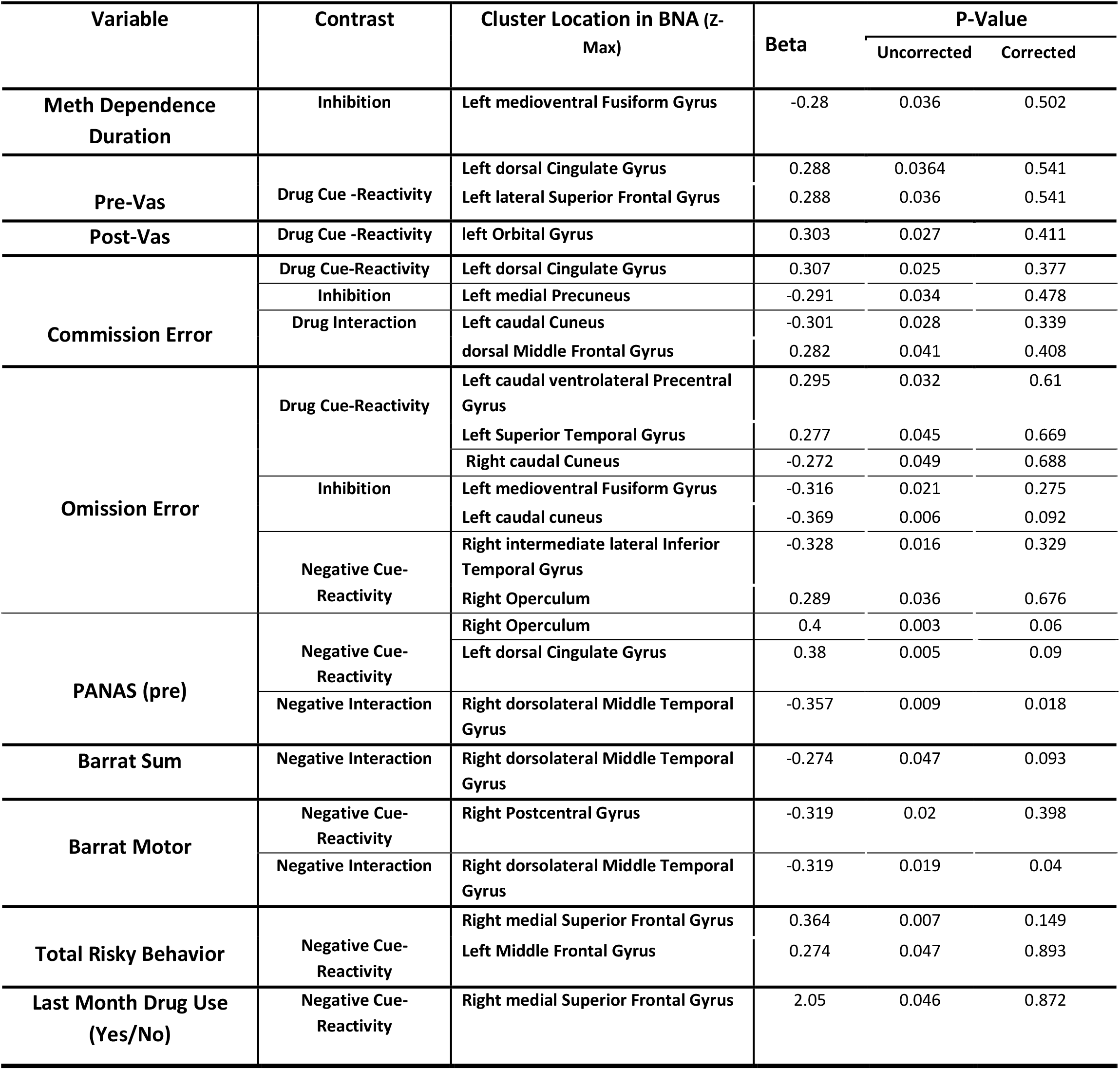
Correlation between demographic and behavioral data with active clusters among three contrasts.

**Figure S1:**
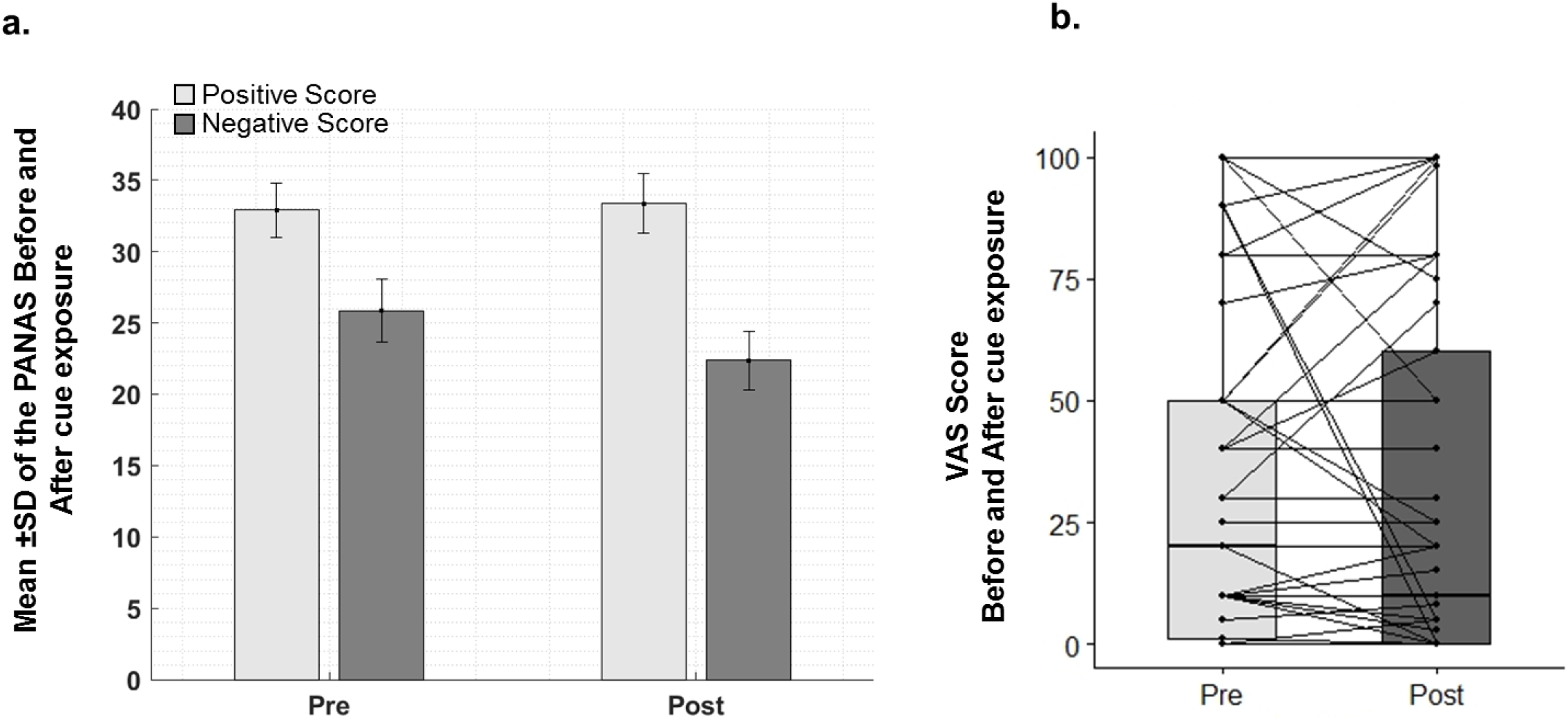
PANAS and VAS before and after scanning.

